# Image-based Explainable Artificial Intelligence Accurately Identifies Myelodysplastic Neoplasms Beyond Conventional Signs of Dysplasia

**DOI:** 10.1101/2025.01.27.25321165

**Authors:** Jan-Niklas Eckardt, Ishan Srivastava, Freya Schulze, Susann Winter, Tim Schmittmann, Sebastian Riechert, Martin M. K. Schneider, Lukas Reichel, Miriam Eva Helena Gediga, Katja Sockel, Anas Shekh Sulaiman, Christoph Röllig, Frank Kroschinsky, Anne-Marie Asemissen, Christian Pohlkamp, Torsten Haferlach, Martin Bornhäuser, Karsten Wendt, Jan Moritz Middeke

## Abstract

Evaluation of bone marrow morphology by experienced hematologists is key in the diagnosis of myeloid neoplasms, especially to detect subtle signs of dysplasia in myelodysplastic neoplasms (MDS). The majority of recently introduced deep learning (DL) models in cytomorphology rely heavily on manually drafted cell-level labels, a time-consuming, laborious process that is prone to substantial inter-observer variability, thereby representing a substantial bottleneck in model development. Instead, we used robust image-level labels for end-to-end DL and trained several state-of-the-art computer vision models on bone marrow smears of 463 patients with MDS, 1301 patients with acute myeloid leukemia (AML), and 236 bone marrow donors. For the binary classifications of MDS vs. donors and MDS vs. AML, we obtained an area-under-the-receiver-operating-characteristic (ROCAUC) of 0.9708 and 0.9945, respectively, in our internal test sets. Results were confirmed in an external validation cohort of 50 MDS patients with corresponding ROCAUC of 0.9823 and 0.98552, respectively. Explainability via occlusion sensitivity mapping showed high network attention on cell nuclei not solely of dysplastic cells. We not only provide a highly accurate model to detect MDS from bone marrow smears, but also underline the capabilities of end-to-end learning to solve the bottleneck of time-consuming cell-level labeling.

## Introduction

Myelodysplastic neoplasms (MDS) encompass clonal myeloid malignancies that are characterized by ineffective hematopoiesis, cytopenia, myelodysplasia, and recurrent genetic events.^1^ The incidence of MDS appears to be underestimated and incidence rates increase dramatically over the age of 70 years (up to an estimated 75:100,000 cases), representing a substantial societal burden in an aging population.^2–4^ Although genetic findings are becoming increasingly important according to the new WHO 2022 classification, accurate cytomorphologic evaluation of the bone marrow remains crucial for the initial diagnosis, response assessment, and detection of disease transformation to acute myeloid leukemia (AML).^5^ While counting myeloblasts is rather straightforward, signs of dysplasia are more subtle and their accurate identification requires experienced investigators. Still, detection is often challenging and prone to inter-observer variability, even for seasoned morphologists^6–8^, and shows discrepancies between site and central review.^9^

In general, cytomorphologic evaluation of bone marrow aspirates in hematology remains essentially unchanged over the last decades, as both preparation and evaluation are performed manually, rendering the entire process time- and cost-intensive, as well as dependent on the experience and subjective judgement of the observer.^10–12^ With the advent of deep learning (DL) systems for computer vision^13^, a multitude of applications in the healthcare sector have been identified where DL is applied in image-based diagnostics.^14,15^ Convolutional neural nets (CNN), which consist of multiple artificial neurons that are interconnected via convoluted deep layers, are commonly used for computer vision tasks.^16^ In cytomorphology, recent studies have utilized neural networks in order to correctly classify peripheral blood and bone marrow cells based on their respective morphology^17–28^, as well as to accurately identify myeloid malignancies.^29,30^

In this study, we used an end-to-end DL system to accurately differentiate between MDS, AML, and healthy donor bone marrow samples based on image-level labels, without the need for manually labeling cells or dysplastic morphologies.

## Methods

### Data sets

We identified 463 MDS patients that have been previously diagnosed and treated at the University Hospital Dresden, Germany. The first control group comprised 1301 AML patients that had been diagnosed and treated under the auspices of the multicenter German Study Alliance Leukemia (SAL) within the following previously reported multicenter trials: AML96^31^ [NCT00180115], AML2003^32^ [NCT00180102], AML60+^33^ [NCT 00180167], and SORAML^34^ [NCT00893373]. Patients were eligible upon diagnosis of MDS or AML according to the revised WHO/ICC criteria^5,35^ age ≥18 years, and available biomaterial at initial diagnosis including bone marrow smears. The second control group consisted of 236 bone marrow samples from healthy bone marrow donors who underwent allogeneic bone marrow donation at our center as previously reported.^36^ An additional external validation cohort was obtained from the Munich Leukemia Laboratory (MLL), Munich, Germany, consisting of 50 patients with diagnosed MDS according to the above-mentioned eligibility criteria. Prior to analysis, written informed consent was obtained from all patients and donors according to the revised Declaration of Helsinki.^37^ All studies were approved by the Institutional Review Board of the TUD Dresden University of Technology (EK 98032010 and EK 289112008).

### Image digitization

Bone marrow smears (BMS) were prepared from anticoagulated bone marrow according to WHO guidelines.^38^ Staining of MDS, AML, and donor BMS was performed with the May-Grünwald-Giemsa method.^11^ Image-level labels were derived from case-level diagnostics, including cytomorphology, histology, cytogenetics and molecular genetics, previously documented for each case during routine diagnostics or as part of the respective clinical trial. Using a Pannoramic 250 FLASH III (3DHISTECH), we obtained high-resolution whole slide images. For every AML patient and bone marrow donor, one image (50x magnification) per whole slide image was obtained using SlideViewer (3DHISTECH). We assumed that subtle signs of dysplasia would not be fully captured in one field of view alone. Therefore, for each MDS patient, we obtained four pictures (50x magnification) of different areas of interest in the BMS.

### Deep learning

#### End-to-end image-level prediction on bone marrow slides

We extended our previously described DL pipeline^29,30^ for binary image-level predictions for the delineation of MDS, AML, and healthy controls. Based on case-level diagnosis, images were labeled with either “MDS”, “AML”, or “healthy donor”. Importantly, no cell-level manual labeling was performed. The pipeline was adapted to evaluate cases in a binary fashion, i.e. MDS vs. AML and MDS vs. healthy donors. Potentially, imbalanced training data can bias a classifier towards the predominant class. Considering the imbalances between the data sets (n=463 samples for MDS with 4 images per patient, resulting in 1852 MDS images in total; n=1301 samples for AML with 1 image per patient; n=236 samples per donor with 1 image per donor), we used image augmentation techniques, such as random sized cropping, color shifting and linear transformations, to balance the data sets for each binary classification task. For all binary classifications, a 5-fold internal cross-validation was used, i.e. a train-test-split of 80:20. Cases that were used for model training were strictly separated from cases that were used for testing. In DL, determination of an optimal model cannot be done a priori, but rather has to be evaluated given the specific use case, data set, and model architecture. Hence, we evaluated six recently introduced DL architectures for computer vision including ResNet-18/34/50/101/152^39^, ResNeXt-50_32×4d/101_32×8d^40^, Wide-ResNet-50/101^41^, DenseNet-121/161/169/201^42^, ShuffleNet v2_x0_5/v2_x1_02^43^, and SqueezeNet v1.1^44^. All DL models were pre-trained on ImageNet data.^45^ The final architecture for each model was determined using automated hyperparameter optimization with the Optuna framework.^46^ DL models were implemented in Python using the PyTorch framework. Computations were performed using the high-performance computing (HPC) cluster of the TUD Dresden University of Technology.

### Performance evaluation

Recall (*syn*.: sensitivity), precision (*syn*.: positive predictive value), and accuracy were used to evaluate classification performances. Recall is defined as the fraction of all positive predictions among all relevant events and precision is defined as the fraction of true positives among all positive predictions. Further, the area-under-the-curve (AUC) was determined for the receiver-operating-characteristic (ROC). All metrics are reported for each binary classification for the internal test sets as well as for the external validation cohort with 95% confidence intervals.

### Explainability of classifications via occlusion sensitivity maps

To highlight network attention and thereby identify morphological cues the network used to delineate MDS, AML, and healthy donors, we used occlusion sensitivity maps (OSM). In OSM, random image areas are iteratively blocked from view of the CNN and classification performance is measured. If the blocked image area is highly relevant for accurate image-level classification, model performance will drop accordingly. This process is repeated for the entire image. Thus, image areas that are crucial for accurate predictions are highlighted so that morphologies that prompt the CNN classifier to predict a label can be evaluated and interpreted.

## Results

### End-to-end deep learning accurately delineates MDS from AML and healthy controls

Baseline characteristics of the MDS patient cohort are shown in Table 1. We evaluated six different neural network architectures^39–44^ for binary classification tasks iteratively. For the distinction between MDS and healthy donors, we found Densenet-201^42^ to provide the highest classification performance, with an accuracy of 0.97791 and a corresponding ROCAUC of 0.9708 (Table 2; Figure 1A). With respect to delineating MDS from AML, the best results were obtained using the Squeezenet^44^ architecture, resulting in an accuracy of 0.98072 and a ROCAUC of 0.9945 (Table 2; Figure 1B). Detailed information on metrics and 95% confidence intervals of the best performing models for each use-case is provided in Table 2. Individual model training on the HPC system for MDS vs. healthy donors and MDS vs. AML took 20 hours each. An external validation set encompassing 50 MDS patients was obtained from the Munich Leukemia Laboratory (MLL). Using our pre-trained models, we achieved an accuracy of 0.9972 with a corresponding ROCAUC of 0.9823 in distinguishing external MDS samples from healthy controls (Table 3, Figure 2A). With respect to delineating external MDS samples from AML, an accuracy of 0.92104 was achieved with a ROCAUC of 0.98552 (Table 3, Figure 2B).

**Figure 1:**
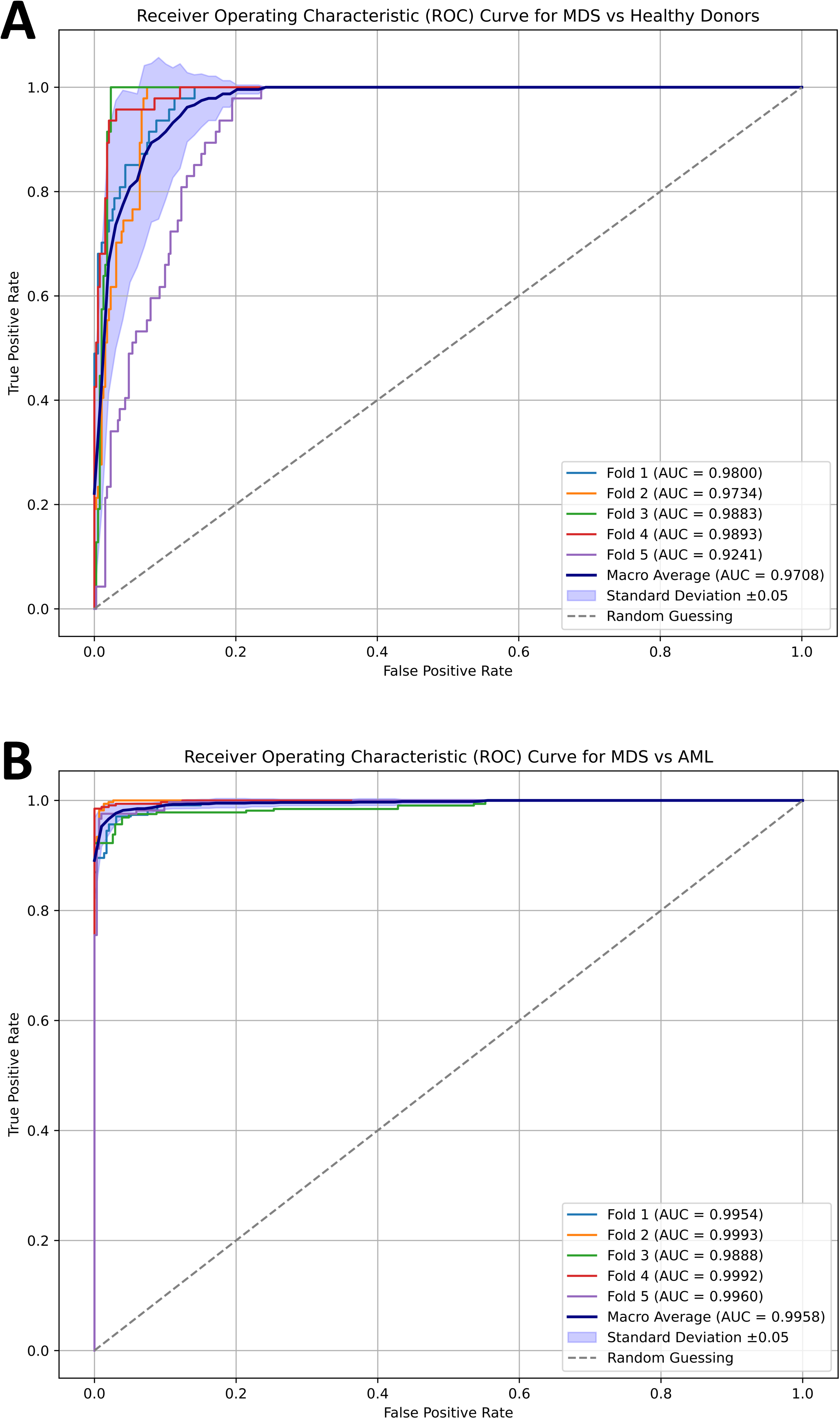
Performance of deep learning models for binary classifications delineating MDS, AML, and healthy donors. The receiver-operating characteristic (ROC) with the corresponding area-under-the-curve (AUC) is depicted for the best performing models for each classification task. For MDS vs. healthy donors, best results were achieved with Densenet-201 (A). For MDS vs. AML, best results were achieved with Squeezenet (B). Internal cross-validation was performed with an 80:20 split. Individual run performance (Fold 1-5; graphs in light blue, orange, green, red, and purple) as well as aggregate macro average performance (graph in dark blue) are reported. Only testing results are reported.

**Figure 2:**
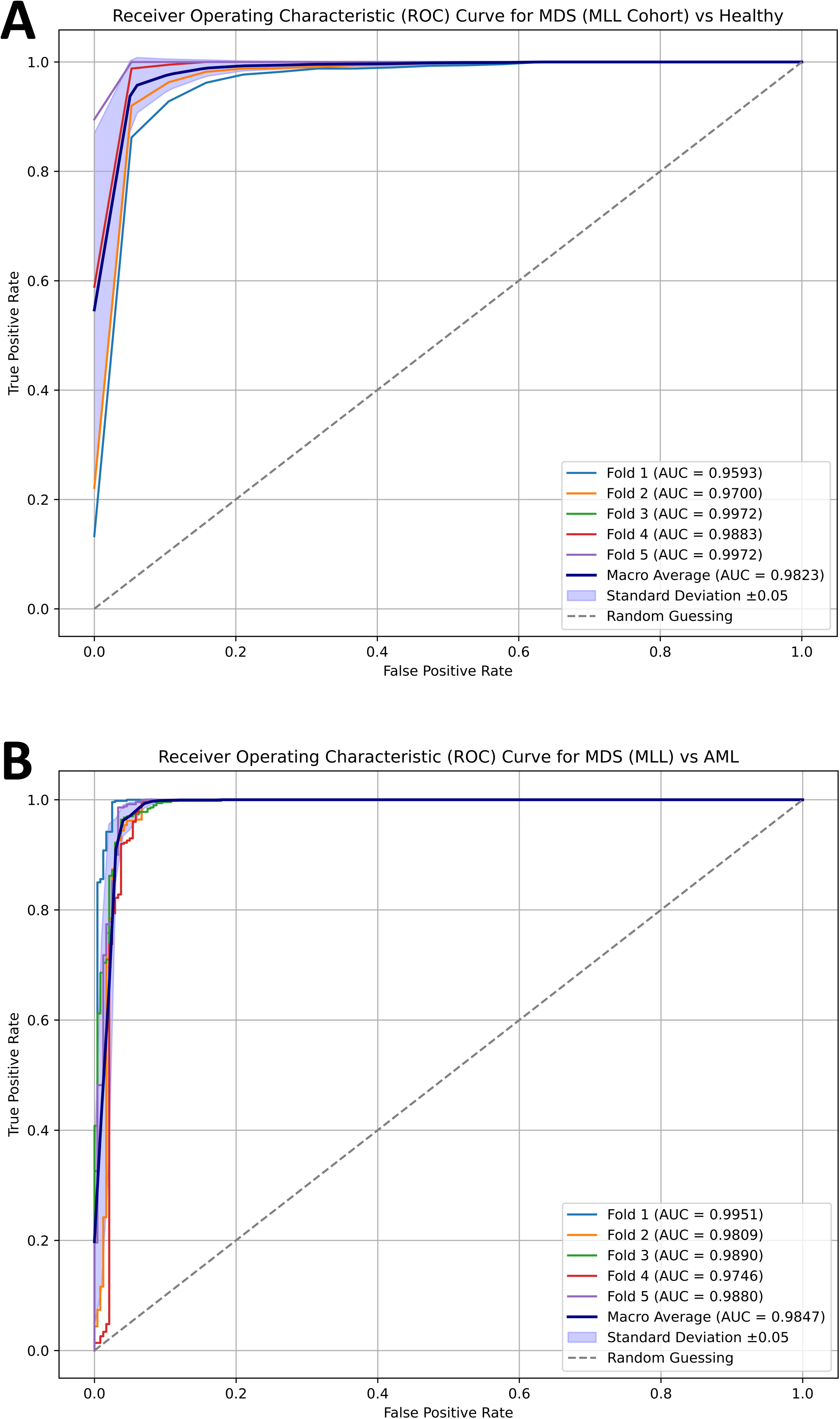
External validation of deep learning models for binary classifications delineating MDS, AML, and healthy donors. The receiver-operating characteristic (ROC) with the corresponding area-under-the-curve (AUC) is depicted for the best performing models for the binary classifications MDS (MLL) vs. healthy donors (A) and MDS (MLL) vs. AML (B). Individual run performance (Fold 1-5; graphs in light blue, orange, green, red, and purple) as well as aggregate macro average performance (graph in dark blue) are reported.

**Table 1.** MDS patient characteristics.

| Parameter |  |
| --- | --- |
| **N** | 463 |
| **Age in years, median (IQR)** | 66 (18-89) |
| **Sex, %** |  |
| Male | 59 |
| Female | 41 |
| **MDS type (WHO 2022), %** |  |
| MDS-5q | 10 |
| with *SF3B1* mutation | 0.1 |
| with *TP53* mutation | 0.1 |
| MDS *biTP53* | 0.2 |
| MDS *SF3B1* | 8.8 |
| MDS-LB | 29.3 |
| MDS, hypoplastic | 2.7 |
| MDS-IB1 | 15.6 |
| MDS-IB2 | 20 |
| MDS with fibrosis | 1.3 |
| MDS/MPN-RS-T | 0.5 |
| CMML-1 | 1.2 |
| CMML-2 | 9.8 |
| **IPSS-R, %** |  |
| Very low risk | 6.4 |
| Low risk | 21.6 |
| Intermediate risk | 38.7 |
| High risk | 22.3 |
| Very high risk | 10.9 |
| **Blood count** |  |
| WBC in GPt/l, median (IQR) | 3.58 (0.57-91.1) |
| Hb in g/dl, median (IQR) | 9.9 (4.4-15.6) |
| Plt in GPt/l, median (IQR) | 96 (3-1531) |
| PB blasts in %, median (IQR) | 0 (0-15) |
| BM blasts in %, median (IQR) | 5.5 (0-26.0) |

*BM* bone marrow, *CMML-1/2* chronic myelomonocytic leukemia subgroup 1/2, *Hb* hemoglobin, *MDS* myelodysplastic neoplasm, *MDS biTP53* MDS with biallelic *TP53* inactivation, *MDS-5q* MDS with low blasts and isolated 5q deletion, *MDS-IB1/2* MDS with increased blasts 1/2, *MDS-LB* MDS with low blasts, *MDS/MPN-RS-T* myelodysplastic/myeloproliferative neoplasm with ring sideroblasts and thrombocytosis, *MDS-SF3B1* MDS with low blasts and *SF3B1* mutation, *N* number, *PB* peripheral blood, *Plt* platelet count, *WBC* white blood cell count.

**Table 2.** Test set performance for binary image-level classifications.

|  | MDS vs. healthy donors |  | MDS vs. AML |  |
| --- | --- | --- | --- | --- |
| DL architecture | Densenet-201 |  | Squeezenet v1.1 |  |
| Accuracy | 0.97791<br>[0.9561 - 0.9977] |  | 0.98072<br>[0.9686 - 0.9904] |  |
|  | MDS | Healthy donors | MDS | AML |
| Precision | 0.9973<br>[0.9948 - 1.0] | 0.8547<br>[0.7121 - 0.9791] | 0.97065<br>[0.9637 - 0.9967] | 0.98118<br>[0.9565 - 0.9904] |
|  | MDS | Healthy donors | MDS | AML |
| Recall | 0.9775<br>[0.9507 - 0.9974] | 0.9787<br>[0.9574 - 1.0] | 0.98180<br>[0.9565 - 0.9906] | 0.98030<br>[0.9616 - 0.9968] |
| ROCAUC | 0.9708<br>[0.9241 - 0.9893] |  | 0.9945<br>[0.98824 - 0.9984] |  |

Brackets indicate 95% confidence intervals. *AML* acute myeloid leukemia, *DL* deep learning, *MDS*
myelodysplastic neoplasm, *ROCAUC* area-under-the-curve of the receiver-operating-characteristic.

**Table 3.** Model performance on external validation set.

|  | **MDS (MLL cohort) vs. healthy donors** |  | **MDS (MLL cohort) vs. AML** |  |
| --- | --- | --- | --- | --- |
| DL architecture | Densenet-201 |  | Squeezenet v1.1 |  |
| Accuracy | 0.9972<br>[0.9811 - 0.9963] |  | 0.92104<br>[0.8905 - 0.9567] |  |
|  | **MDS** | **Healthy** | **MDS** | **AML** |
| Precision | 0.9925<br>[0.9892 - 1.0] | 0.9852<br>[0.9787 - 0.9957] | 0.91418<br>[0.8880 - 0.9398] | 0.94668<br>[0.8245 - 1.0] |
|  | **MDS** | **Healthy** | **MDS** | **AML** |
| Recall | 0.9970<br>[0.9940 - 0.9980] | 0.9938<br>[0.9755 - 1.0] | 0.97516<br>[0.9139 - 1.0] | 0.80834<br>[0.7375 - 0.8667] |
| ROCAUC | 0.9823<br>[0.9593 - 0.9972] |  | 0.98552<br>[0.9746 - 0.9951] |  |

Brackets indicate 95% confidence intervals. *AML* acute myeloid leukemia, *MDS* myelodysplastic neoplasm, *MLL* Munich Leukemia Laboratory, *ROCAUC* area-under-the-curve of the receiver-operating-characteristic.

### Explainable predictions via occlusion sensitivity maps

In order to make results interpretable to cytomorphologists, we used OSM that iteratively blocks image areas from neural network evaluation and thus highlights (in red) image areas that are of high importance for accurate predictions. In a proof-of-concept fashion, we found OSM to be cell-specific, indicating that network attention is being focused on cells, rather than background or smudge. Network attention was focused on cells in granulopoiesis and erythropoiesis and on megakaryocytes (Figure 3). Interestingly, neural networks focused not only on signs of dysplasia, but also on cells we deemed morphologically inconspicuous. High attention was given to defined signs of dysplasia involving altered nuclear morphology such as chromatin clumping, dysfunctional segmentation, or double nuclei. However, at times, high network attention was also given to cells with no apparent dysplasia as per conventional definition,^11,47^ while network attention in these cells was also mainly focused on the nucleus, sometimes including the perinuclear zone. This indicates more intricate and subtle morphological alterations unquantifiable by human observers. However, other signs of dysplasia, such as hypogranulation, were disregarded by our model. This could possibly be either due to confidence saturation - meaning the model found enough reasons in a given field of view to confidently predict MDS without paying attention to all apparent signs of dysplasia (defined or not) - or low-ranking signs of dysplasia that were not learned in the training process due to their limited weight in making accurate predictions.

**Figure 3:**
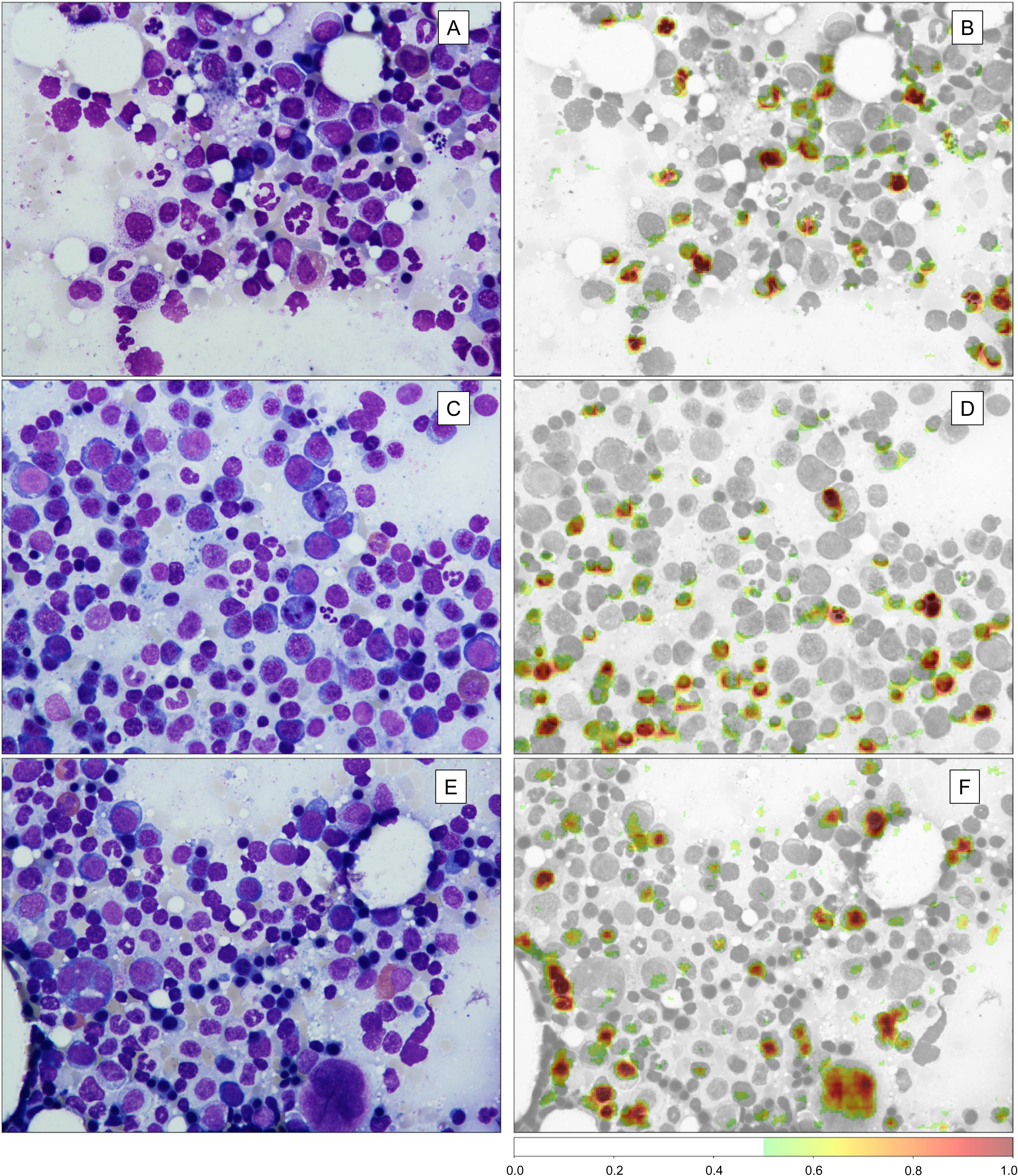
Occlusion Sensitivity Mapping (OSM) highlights network attention for explainable output interpretation. OSM iteratively blocks image areas from being evaluated by the deep learning network. If an image area is highly important for classification, the network’s performance will thus drop substantially in the given iteration. Image areas that are of high importance for correct classification can thereby be highlighted (high attention shown in red). A standard field of view of bone marrow smears from MDS patients is shown in A, C, and E. The corresponding OSM is displayed in B, D, and F, respectively. First, in a proof-of-concept fashion, the network focuses its attention on cells and specifically on nuclei. It does not consider background, noise or smudge as important for classification. Second, high attention is directed at erythropoietic and granulopoietic cells as well as megakaryocytes.

## Discussion

Using end-to-end DL, we developed a software framework to distinguish between MDS, AML, and healthy controls with very high accuracy based on BMS from 2000 individual patients and bone marrow donors. Importantly, we have demonstrated that information abstraction even in MDS with often subtle morphologies is feasible using end-to-end learning, in contrast to recent studies in hematology that primarily rely on the generation of cell-level labels.^17–28,48,49^ Using the latter approach, a bottom-up system has to be devised where first thousands (usually hundreds of thousands) of labels are required to build a robust classifier, and second individual cell-level predictions have to be aggregated to generate a diagnosis-level prediction. Apart from being obviously time-consuming and cost-ineffective, the generation of cell-level labels, i.e. the ground truth many classifiers in hematology currently are based on, is flawed due to substantial classification biases. For instance, Sasada et al.^7^ evaluated divergence in cell classifications on 100,000 hematopoietic cells of 499 MDS patients. Up to eleven experienced observers evaluated each cell image, however, only 55.6% of classifications were found to match and especially low classification overlap was reported for dysplastic morphologies like hypo-granularity and Pseudo-Pelger-Huët anomaly.^7^ This bottleneck and pitfall of cell-level labeling can essentially be bypassed by an end-to-end approach, such as ours. Our ground truth labels are not derived from subjective observer judgment. Instead, they are established by routine diagnostics, including cytomorphology, histology, flow cytometry, cytogenetics, molecular genetics and clinical examination, which provide image-level ground truth labels that are much more robust.

With respect to explainability, DL is often referred to as a ‘black box’.^50^ The often elusive decision-making of neural networks substantially hampers interpretability and thus acceptance of DL models in such high-risk applications as cancer diagnostics. Using OSM (among other methods of explainability^51^) not only enables internal proof-of-concept, but also provides additional information to the human observer, as novel features that are important for prediction can be investigated that otherwise would elude the human eye. Interestingly, our classifiers showed high attention for nuclei not only of dysplastic cells, but also for cells that we did not deem to be morphologically suspicious for dysplasia. Potentially, this alludes to a digital biomarker in MDS distinct from classical signs of dysplasia. Future work will focus on correlating attention maps with genetic alterations and/or gene expression in MDS. While certain molecular alterations have already been linked to certain morphologies, such as mutated *SF3B1* in MDS with ringsideroblasts^52,53^, CNNs can potentially be used to identify novel gene-morphology links. For instance, Brück et al.^54^ used CNNs on MDS bone marrow core biopsies to predict mutations of *TET2*, spliceosome genes and monosomy 7. Further, Nagata et al.^55^ previously demonstrated a link between MDS morphology (assessed by pathologists) and genomic profiles. This suggests a starting point for CNNs to link gene alterations with specific morphologies and may potentially lead to an image-based predictor of genetic profiles and consequentially patient risk and outcome.

Our study is limited by several factors. As is the case for most recent studies of computer vision in (hemato-)pathology, our analysis is based on retrospective data. While external validation confirmed high classification accuracy, prospective validation is still warranted. In our study, we differentiated only between AML, MDS, and healthy bone marrow donors in a binary way. Still, some dysplastic morphologies can also be present to a certain degree in non-malignant disorders^56^ such as congenital syndromes^57^, nutritional deficiencies^58,59^, infectious disease^60^, and drug- or toxin-mediated bone marrow damage^61,62^. To increase routine applicability of our DL framework, future work will also focus on acquiring image data from reactive and non-neoplastic specimen exhibiting bone marrow dysplasia in order to make our classifier more versatile and applicable in clinical routine.

In summary, we have developed a DL framework trained on patient and donor samples, achieving high accuracies in our internal test set and external validation set in distinguishing between MDS, AML, and healthy bone marrow donors.

## Data Availability

All data produced in the present study are available upon reasonable request to the authors.

## List of Abbreviations

AML: acute myeloid leukemia
BMS: bone marrow smear(s)
CNN: convolutional neural net
DL: deep learning
MDS: myelodysplastic neoplasm(s)

## Acknowledgements

The authors are grateful to the Centre for Information Services and High-Performance Computing of the TUD Dresden University of Technology for providing its facilities for training and execution of deep learning models. This study was funded in part by Novartis Oncology. The funder had no role in conceptualization, design, data collection, analysis, decision to publish, or preparation of the manuscript.

## Author Contributions

J-NE and JMM conceptualized the project. J-NE, FS, MEVG, KS, ASS, CR, UP, CP, TH, MB, and JMM provided patient samples. J-NE, FS, MS, LR, and MEHG acquired BMS images. IS, TS, SR, and KW developed computer vision models. All authors analyzed and interpreted the data. J-NE wrote the initial draft. All authors approved the final version of the manuscript and agreed to be accountable for all aspects of the work.

